# A cluster-randomized trial of client and provider directed financial interventions to align incentives with appropriate case management in private medicine retailers: results of the TESTsmART Trial in Lagos, Nigeria

**DOI:** 10.1101/2024.01.30.24302026

**Authors:** T. Visser, J. Laktabai, E. Kimachas, J. Kipkoech, D. Menya, D. Arthur, Y. Zhou, T. Chepkwony, L. Abel, E. Robie, M. Amunga, G. Ambani, P. Uhomoibhi, N. Ogbulafor, B. Oshinowo, O. Ogunsola, M. Woldeghebriel, E. Garber, T. Olaleye, N. Eze, L. Nwidae, P. Mudabai, J.A. Gallis, C. Fashanu, I. Saran, A. Woolsey, E.L. Turner, W. Prudhomme O’Meara

## Abstract

Malaria remains a major health priority in Nigeria. Among children with fever who seek care, less than a quarter gets tested for malaria, leading to inappropriate use of the recommended treatment for malaria; Artemether Combination Therapies (ACT). Here we test an innovative strategy to target ACT subsidies to clients seeking care in Nigeria’s private retail health sector who have a confirmed malaria diagnosis. We supported point-of-care malaria testing (mRDTs) in 48 Private Medicine Retailers (PMRs) in the city of Lagos, Nigeria and randomized them to two study arms; a control arm offering subsidized mRDT testing for USD $0.66, and an intervention arm where, in addition to access to subsidized testing as in the control arm, clients who received a positive mRDT at the PMR were eligible for a free (fully subsidized) first-line ACT and PMRs received USD $0.2 for every mRDT performed. Our primary outcome was the proportion of ACTs dispensed to individuals with a positive diagnostic test. Secondary outcomes included proportion of clients who were tested at the PMR and adherence to diagnostic test results. Overall, 23% of clients chose to test at the PMR. Test results seemed to inform treatment decisions and resulted in enhanced targeting of ACTs to confirmed malaria cases with only 26% of test-negative clients purchasing an ACT compared to 58% of untested clients. However, the intervention did not offer further improvements, compared to the control arm, in testing rates or dispensing of ACTs to test-positive clients. We found that ACT subsidies were not passed on to clients testing positive in the intervention arm. We conclude that RDTs could reduce ACT overconsumption in Nigeria’s private retail health sector, but PMR-oriented incentive structures are difficult to implement and may need to be complemented with interventions targeting clients of PMRs to increase test uptake and adherence. Clinical Trials Registration Number: NCT04428307

## Introduction

Malaria remains an urgent health priority in Nigeria where it is a leading cause of mortality. Globally, Nigeria accounts for 27% of all malaria cases, exceeding 66 million per year [1]. Reported cases, which are already the highest in the world, are on the rise, suggesting that beyond an expansion of malaria testing and improved surveillance, the malaria burden might be getting worse [2]. Recent survey results show that among children under 5 who had a fever in the two weeks before the survey, only 24% had blood taken from a finger or heel for malaria testing [3]. Private health providers including private medicine retailers (PMRs) such as Patent and Proprietary Medicine Vendors (PPMVs) and Community Pharmacies play an important role in providing basic health services in Nigeria, including management of suspected malaria cases. PMRs are widespread across Nigeria and are often the first point of care for febrile patients. Household survey data suggests up to 60% of febrile children first seek care in PMRs [3]. Compliance with national malaria treatment guidelines is generally considered poor in these outlets [4,5,6]. Few clients receive a confirmatory malaria diagnosis prior to treatment, with PMRs and their clients often forgoing the test or taking antimalarials even when the test result is negative [7].

Here we report the results of the *Malaria diagnostic testing and conditional subsidies to target ACTs in the retail sector* (TESTsmART) trial in Nigeria, a two-arm cluster randomized trial designed to test the effect of conditional ACT subsidies and modest provider incentives on appropriate use of ACT and malaria Rapid Diagnostic Test (mRDT) in the retail sector. The study was conducted in the urban center of Lagos State, located in South-Western region of Nigeria, where 2.6 million malaria cases (3.8% of Nigeria’s total cases) were recorded in 2021 [2].

## Methods

### Overall study design

This study was a two-arm cluster randomized controlled trial focused on malaria diagnostic testing and conditional ACT subsidies with the goal of evaluating the effect of provider-directed and client-directed interventions on improving the management of suspected malarial fevers that receive care in the retail sector. The study site was the urban center of Lagos, Lagos State, Nigeria. We enrolled 48 PPMV outlets (clusters) in the study. Inclusion criteria for PPMV outlets were: (i) Routinely stock and sell ACTs; (ii) Willing to acquire mRDTs and use in diagnosing malaria for clients; (iii) Willing to use a phone/app to collect/report data and receive incentives to conduct mRDTs; (iv) Willing to allow a data collector to conduct patient exit interviews for several days each month at the outlet; (v) Up-to-date license/registration. Exclusion criteria for outlets were (i) Having challenges with network connectivity at the outlet; (ii) Participating in other studies/NGO projects and (iii) Having any agreements with drug/diagnostic marketers.

The diagnosis and treatment choices made during each transaction were captured using a mobile phone reporting app. This app was designed specifically for the purposes of the study. Each outlet was provided a mobile phone with the app and monthly data bundles to use the app. Outlet attendants were asked to use the app for each client with suspected malaria. The app included a few questions on the client’s symptoms and if an RDT was conducted, it prompted the user to take and upload a picture of the RDT result.

Study outcomes were collected through exit interviews with clients, who sought care for febrile illness at any of the enrolled PPMVs. Inclusion criteria for clients to be eligible for the exit interview were: (i) Participants had a fever or history of fever in the last 48 hours or malaria-like illness; (ii) Individuals with malaria-like illness was present at recruitment and (iii) Older than one year of age. Exclusion criteria were (i) Individuals with any signs of severe illness requiring immediate referral; (ii) Individuals who had taken an antimalarial in the last seven days, including for the current illness; (iii) Individuals that were under 18 years of age without a parent or legal guardian present and (iv) Individuals that were unable to consent. Written informed consent was obtained from the parent/guardian of each participant under 18 years of age.

This study is linked to a similar cluster randomized controlled trial in the contrasting rural study site of the region around Webuye in western Kenya. These results are pending publication elsewhere [8]. Full details of the study design were previously reported in Woolsey *et al* [9, 10].

### Study area

The study was conducted in Lagos, Nigeria, a large urban metropolis in southern Nigeria where malaria prevalence is around 3% and point-of-care rapid testing through mRDTs has been permitted in PPMVs since 2011 [11].

### Intervention

The two intervention arms considered for evaluation were:

1. Control intervention: mRDTs were made available at a wholesale price to the participating PPMV (120 Naira or approximately USD $0.31 per test), as per the USD/Naira exchange rate at the onset of the implementation phase in March 2022. Outlet owner/attendants were trained to use the mobile reporting app. mRDTs were offered to clients at a pre-determined price (250 Naira per test or approximately USD $0.66).
2. Provider-directed and client-directed intervention: in addition to the interventions implemented in the control outlets, the retail outlet owner received a small incentive (250 Naira or approximately USD $0.66) for each mRDT they conducted and reported using the mobile app. Clients visiting outlets in this arm received a free ACT (cost equivalent to 850 Naira or approximately USD $2.12 for adults and 650 Naira or approximately USD $1.62 for children) if they purchased an mRDT and received a positive test result (conditional subsidy).

Originally, the study was planned as a four-arm study, splitting provider directed and client directed interventions in separate arms in addition to a control and the combined intervention arm, but due to substantial differences between the initial study assumptions and actual observations during the first seven months of data collection, the study was re-started in February 2022 with two arms rather than four to address the anticipated loss of power. The recruitment period for the 2-arm design started March 1^st^, 2022, and ran until February 28^th^, 2023. For a detailed explanation of the change from the originally planned 4-arm design to a 2-arm design please refer to Woolsey *et al* [10].

### Study outcome measures

The primary outcome was ACT consumption by parasitologically confirmed malaria cases, defined as the proportion of ACTs that were dispensed to malaria test-positive clients, including those who were tested at the outlet and those who came with documentation of a positive test result (either microscopy or mRDT) from another provider. There were four secondary outcomes, all binary:

1. Use of malaria rapid diagnostic test: Proportion of suspected malaria cases that received a malaria test, where a suspected case was any client who was tested with an mRDT or was untested but purchased an antimalarial (AM) of any type.
2. Adherence to mRDT result: Proportion of malaria tested clients whose treatment adhered to test results (tested positive with mRDT and purchased ACT or tested negative with mRDT and did not purchase AM).
3. Appropriate case management: Proportion of suspected malaria cases (defined as above in 1.) that were managed appropriately (same numerator as for “Adherence to mRDT result” outcome, but with all suspected malaria cases as denominator).
4. ACT use among the untested: Proportion of untested clients who purchased ACT.

### Power calculations

All power calculations were based on pairwise comparison of two proportions using a formula for comparing two proportions under a cluster-randomized trial design [12]. We estimated the intra-class correlation coefficient (ICC) for the primary outcome to be 0.037. In order to account for varying cluster sizes, we modified the Hayes and Moulton formula for comparing two proportions by replacing the cluster size (m) with *m*/(1 + *CV*^2^), where *CV* is the coefficient of variation of cluster sizes and was estimated to be 0.72 (13). Based on these assumptions, we expected to achieve greater than 90% power to detect a difference of 12 percentage points in the primary outcome for our comparison of the intervention arm to the control arm.

### Randomization and recruitment

For the purposes of the two-arm trial, 48 retail PPMVs were equally randomly assigned to each of the two arms by the study statistician. Randomization was constrained to stratify by three geographic regions of Lagos, and additionally constrained such that any outlets in close proximity to one another (<0.5km) were assigned to the same arm to avoid potential contamination of treatment effects. For additional details about the randomization procedures before and after the design change from four arms to two arms, see the analysis plan in the supplement.

### Study implementation

Enrolled PPMV outlets were trained on Nigeria’s national malaria case management guidelines, correct mRDT procedures, and the use of the study mobile app to record client encounter data and take photos of mRDT results. Outlets were also equipped with arm specific leaflets and banners placed outside of the outlet, encouraging customers to test for malaria in the outlet. During a 3-month pilot period, the outlets started collecting data on the mobile app and performing mRDTs. Outlets were monitored closely to make sure testing and app issues were resolved during the pilot period. After the pilot period, outlets were trained in arm-specific groups on the intervention guidelines for client and provider-directed incentives, as applicable. Outlets were also informed of the exit interview process being conducted outside of their outlet, independently of the client-outlet interaction.

Throughout the 12-month intervention period, the study team conducted regular supervision visits to ensure good adherence to testing and safety procedures, troubleshoot challenges with the mobile app and provide onsite mentorship to all outlets. Outlets that were running low on RDTs were urged to contact the supplier contracted to supply mRDTs for deliveries directly to the outlet. Any abnormal mRDT photographs observed submitted via the app were passed along to supervisors to discuss with the outlets.

Subsidies and incentives were paid weekly to bank accounts of the owners of the intervention outlets. Outlets allocated to the intervention arm agreed to give free Artemether Lumefantrine (AL) to test-positive clients for which they were reimbursed by the study (650 Naira for children 9 years and below and 850 Naira for clients 10 years and above, correlating with the price of the different dosages for each age range). Outlets also received a small payment for administering mRDTs to clients (100 Naira), regardless of test result. Payments were calculated based on photographs of mRDTs uploaded to the study app. Outlets were paid for the total number of tests conducted, and for the total number of positive mRDTs, as observed through the study app. For this study, reimbursable ACTs were restricted to authorized AL brands (i.e., Registered by the national regulatory authority in Nigeria; the National Agency for Food and Drug Administration and Control (NAFDAC) and approved by World Health Organization Pre-Qualification Program) in a tablet formulation we found to be widely available among the participating outlets (Lonart/ CoArtem/ Lumartem).

### Data collection

Interviews were conducted with clients departing from participating PPMVs (study clusters) on random days of the week. All clients exiting the PPMV that day were eligible to be screened. Exit interviewers were instructed to make no pre-judgements about clients but rather approach each client exiting the outlet. Data for participant exit interviews was collected electronically via tablet. The primary tool for developing the data collection forms was REDCap hosted at Duke University.

### Statistical analysis

Characteristics of enrolled participants were summarized by study arm using means and standard deviations for continuous variables and counts and percentages for categorical variables. We analyzed all client-level self-reported primary and secondary outcomes using a modified Poisson approach with robust standard errors, with log link to estimate risk ratios (RRs) and identity link to estimate risk differences (RDs) [14,15]. The modified Poisson approach was implemented within the generalized estimating equations (GEE) framework to account for clustering of outcomes by outlet and utilizing an independence working correlation to avoid bias in estimating the treatment effect [16]. Moreover, because there were fewer than 50 clusters, the robust standard errors were further adjusted for potential “small-sample” bias using the Kauermann-Carroll correction [17, 18]. All models included fixed effects for the intervention arm indicators, the stratification variable (county), and time (in months, including linear, quadratic, and cubic terms) to account for potential imbalanced recruitment over the 12-month follow-up period (minimally adjusted models). Model-based ICC for the primary outcome was calculated using this minimally adjusted model. We also present fully adjusted models which include the following potential confounder variables: client gender, client age, level of schooling, and wealth index.

We intended to estimate correlation parameters using the matrix-adjusted estimating equation (MAEE) approach [19], but this approach did not converge. Therefore, we reverted to the GEE approach with method of moments estimation of correlation parameters. Inference for parameters of the mean model was based on the t-statistic with degrees of freedom *I* ― *p*, where *I* was the number of clusters (PPMV outlets) and *p* was the number of parameters in the mean model (specifically of the cluster-level covariates, including the intercept, treatment indicator, and time). All analyses were based on the intention-to-treat principle. Since we did not have longitudinal follow-up of clients, we did not need to account for missing data due to attrition of clients. Clients missing specific data elements were excluded from models requiring those variables as indicated in the tables. Overall, there was relatively little missing data for variables included in the minimally adjusted models. For the fully adjusted models, approximately 15% of participants were missing data required for the construction of the wealth score, and 10% were missing information on their education level.

### Ethical considerations and trial registration

The study was reviewed and approved by CHAI’s Scientific and Ethical Review Committee (SERC) on October 22^nd^ 2019; Duke University Institutional Review Board (Pro00104256) and the Health Research and Ethics Committee Secretariat (HREC) at the Lagos State University Teaching Hospital (LASUTH) (LREC/06/10/1304) in Lagos, Nigeria. The study is registered in ClinicalTrials.gov (NCT04428307) and the protocol and revisions to the original design were published in advance [9,10].

## Results

### Population analysis

Between early March 2022 and late February 2023, data were collected from exiting clients on 251 outlet-days of observation. Out of 12,947 clients screened, 2,441 or 19% were eligible (i.e., met the inclusion criteria) and provided consent across the two arms. Most ineligible clients (8,608 or 82%) indicated not having a fever or malaria-like illness; some (1,049 or 10%) ineligible clients indicated that they visited the outlet for someone else with malaria-like symptoms that was not present. Of the 2,205 enrolled, consenting and eligible clients, 1,815 or 82% were adults who had malaria-like symptoms; 388 or 18%, were the caregiver of a sick child. The median age of the child was 5. Eligible clients showed an even split in gender. Almost half (49% or 1,065) of respondents (adult client or caregiver of child client) were between ages 26-39, and 17% or 367 respondents were between ages 18-25. Over the 12-month collection period starting in early March 2022, 31% of exit interviews were collected between September and November 2022, with the other interviews split evenly across the remaining three 3-month periods (20-25%). Almost half of respondents (915 or 47%) achieved high school as the highest level of education, 451 or 23% a university degree and 404 or 21% a vocational degree. We did not observe any notable differences in respondent characteristics across arms (Fig 1, Table 1).

**Figure 1:**
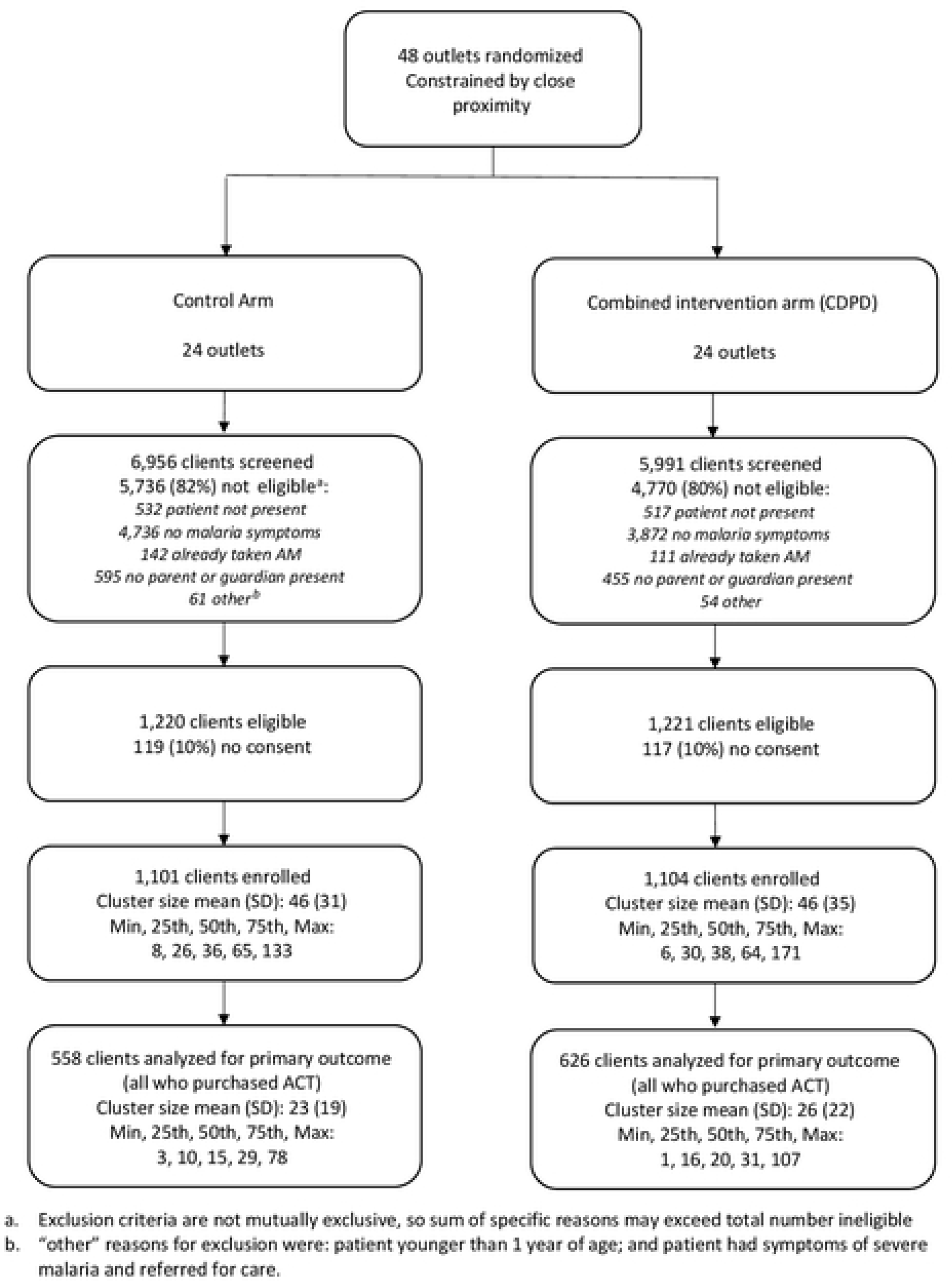
Flow diagram of shop enrollment, randomization and client Interviews.

**Table 1:**
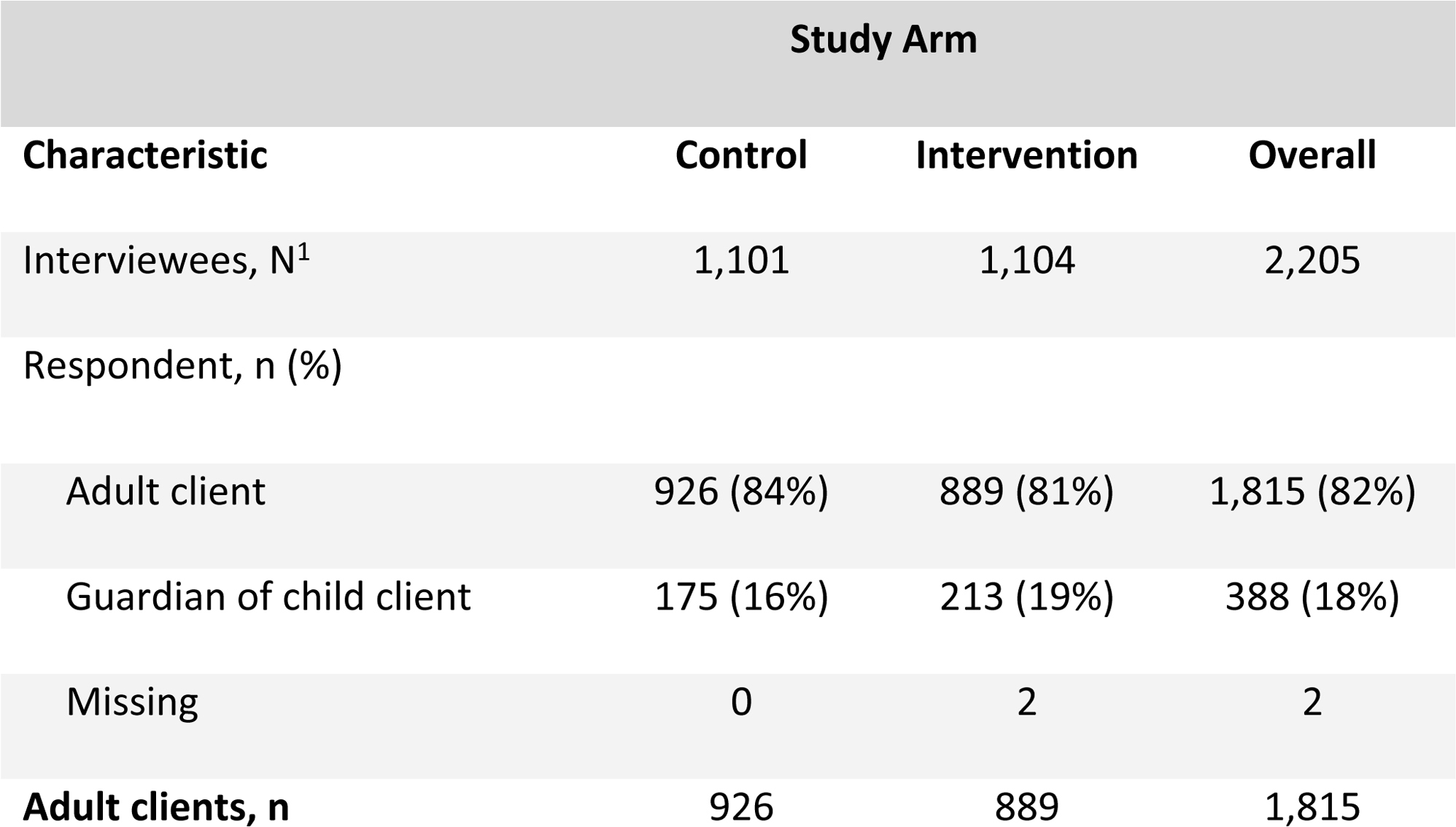
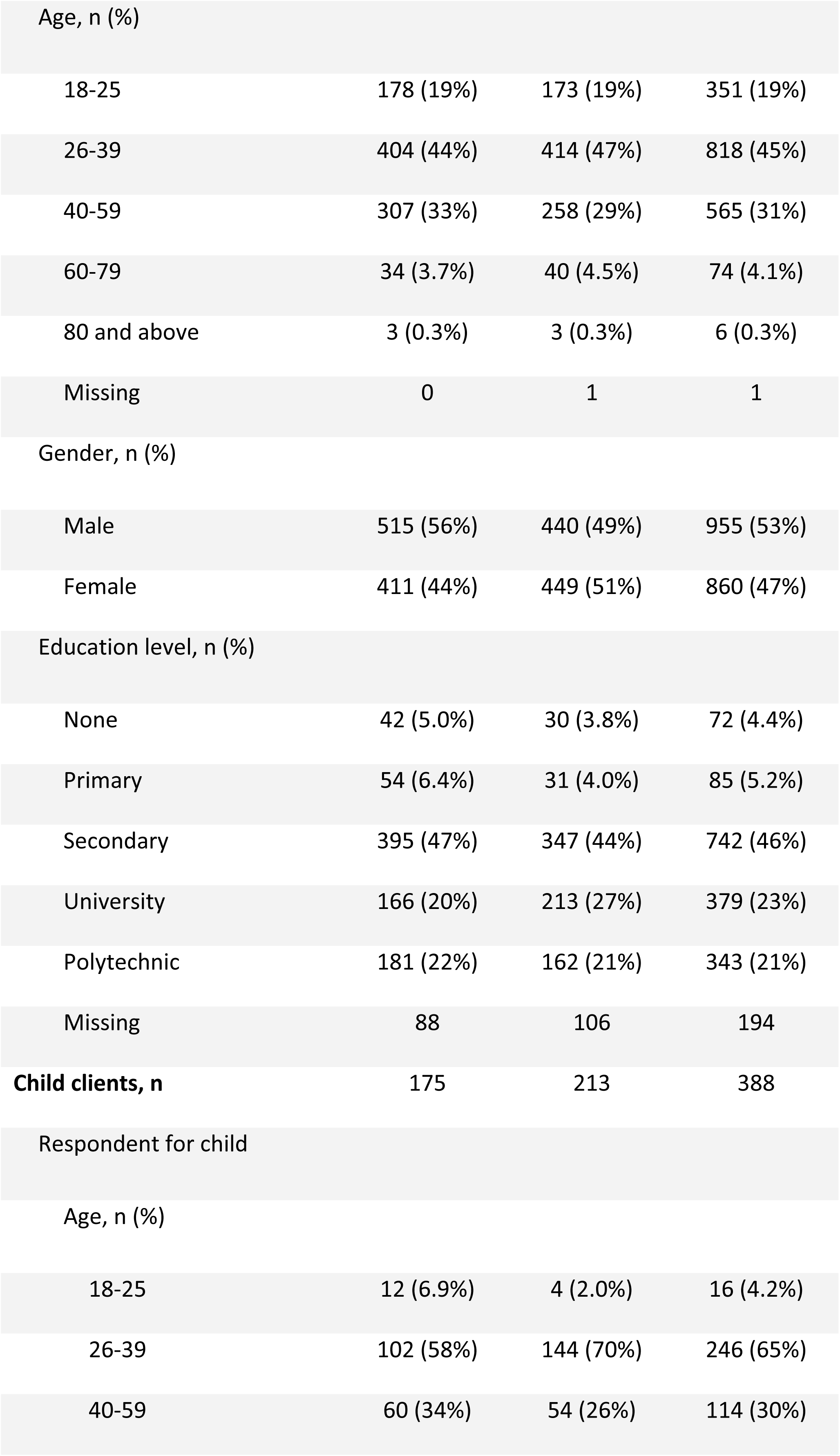
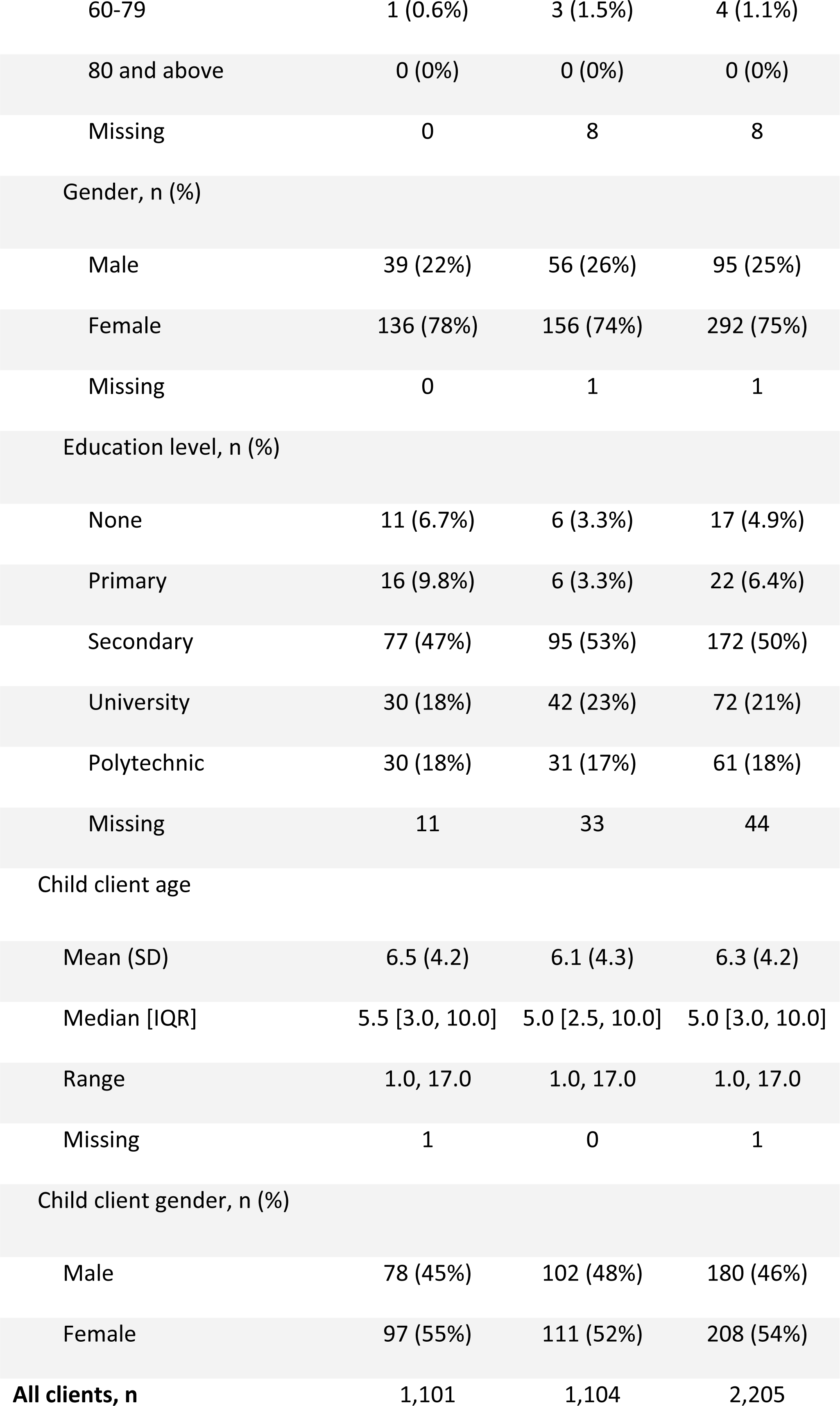
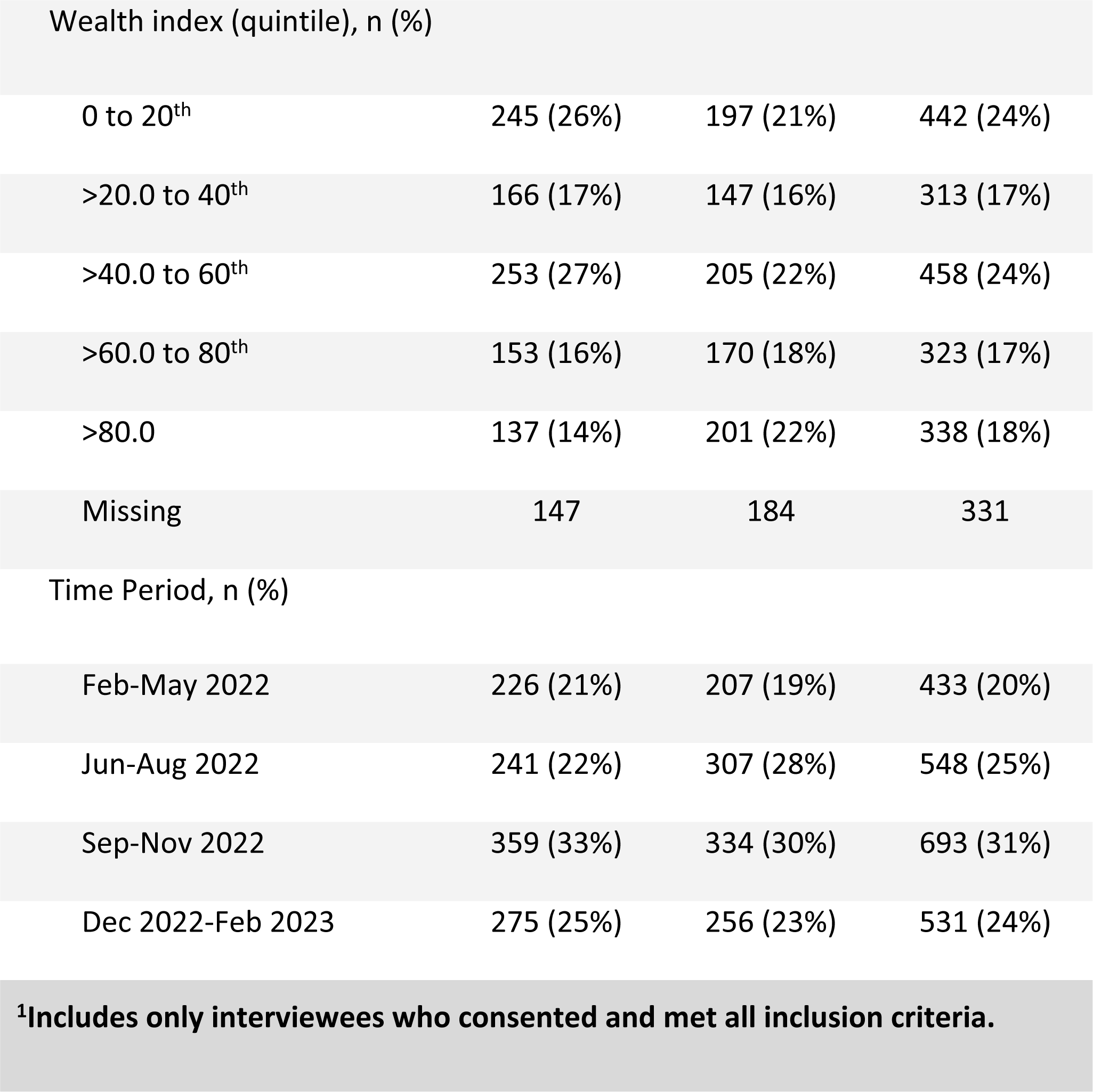
Characteristics of participants enrolled in TESTsmART by arm.

| Characteristic | Study Arm |  |  |
| --- | --- | --- | --- |
|  | Control | Intervention | Overall |
| Interviewees, N<sup>1</sup> | 1,101 | 1,104 | 2,205 |
| Respondent, n (%) |  |  |  |
| Adult client | 926 (84%) | 889 (81%) | 1,815 (82%) |
| Guardian of child client | 175 (16%) | 213 (19%) | 388 (18%) |
| Missing | 0 | 2 | 2 |
| **Adult clients, n** | **926** | **889** | **1,815** |

| Study Arm |  |  |  |
| --- | --- | --- | --- |
| Characteristic | Control | Intervention | Overall |
| Age, n (%) |  |  |  |
| 18-25 | 178 (19%) | 173 (19%) | 351 (19%) |
| 26-39 | 404 (44%) | 414 (47%) | 818 (45%) |
| 40-59 | 307 (33%) | 258 (29%) | 565 (31%) |
| 60-79 | 34 (3.7%) | 40 (4.5%) | 74 (4.1%) |
| 80 and above | 3 (0.3%) | 3 (0.3%) | 6 (0.3%) |
| Missing | 0 | 1 | 1 |
| Gender, n (%) |  |  |  |
| Male | 515 (56%) | 440 (49%) | 955 (53%) |
| Female | 411 (44%) | 449 (51%) | 860 (47%) |
| Education level, n (%) |  |  |  |
| None | 42 (5.0%) | 30 (3.8%) | 72 (4.4%) |
| Primary | 54 (6.4%) | 31 (4.0%) | 85 (5.2%) |
| Secondary | 395 (47%) | 347 (44%) | 742 (46%) |
| University | 166 (20%) | 213 (27%) | 379 (23%) |
| Polytechnic | 181 (22%) | 162 (21%) | 343 (21%) |
| Missing | 88 | 106 | 194 |
| **Child clients, n** | **175** | **213** | **388** |
| Respondent for child |  |  |  |
| Age, n (%) |  |  |  |
| 18-25 | 12 (6.9%) | 4 (2.0%) | 16 (4.2%) |
| 26-39 | 102 (58%) | 144 (70%) | 246 (65%) |
| 40-59 | 60 (34%) | 54 (26%) | 114 (30%) |
|  | Control | Intervention | Overall |
| 60-79 | 1 (0.6%) | 3 (1.5%) | 4 (1.1%) |
| 80 and above | 0 (0%) | 0 (0%) | 0 (0%) |
| Missing | 0 | 8 | 8 |
| Gender, n (%) |  |  |  |
| Male | 39 (22%) | 56 (26%) | 95 (25%) |
| Female | 136 (78%) | 156 (74%) | 292 (75%) |
| Missing | 0 | 1 | 1 |
| Education level, n (%) |  |  |  |
| None | 11 (6.7%) | 6 (3.3%) | 17 (4.9%) |
| Primary | 16 (9.8%) | 6 (3.3%) | 22 (6.4%) |
| Secondary | 77 (47%) | 95 (53%) | 172 (50%) |
| University | 30 (18%) | 42 (23%) | 72 (21%) |
| Polytechnic | 30 (18%) | 31 (17%) | 61 (18%) |
| Missing | 11 | 33 | 44 |
| Child client age |  |  |  |
| Mean (SD) | 6.5 (4.2) | 6.1 (4.3) | 6.3 (4.2) |
| Median [IQR] | 5.5 [3.0, 10.0] | 5.0 [2.5, 10.0] | 5.0 [3.0, 10.0] |
| Range | 1.0, 17.0 | 1.0, 17.0 | 1.0, 17.0 |
| Missing | 1 | 0 | 1 |
| Child client gender, n (%) |  |  |  |
| Male | 78 (45%) | 102 (48%) | 180 (46%) |
| Female | 97 (55%) | 111 (52%) | 208 (54%) |
| **All clients, n** | **1,101** | **1,104** | **2,205** |
|  | Control | Intervention | Overall |
| Wealth index (quintile), n (%) |  |  |  |
| 0 to 20<sup>th</sup> | 245 (26%) | 197 (21%) | 442 (24%) |
| >20.0 to 40<sup>th</sup> | 166 (17%) | 147 (16%) | 313 (17%) |
| >40.0 to 60<sup>th</sup> | 253 (27%) | 205 (22%) | 458 (24%) |
| >60.0 to 80<sup>th</sup> | 153 (16%) | 170 (18%) | 323 (17%) |
| >80.0 | 137 (14%) | 201 (22%) | 338 (18%) |
| Missing | 147 | 184 | 331 |
| Time Period, n (%) |  |  |  |
| Feb-May 2022 | 226 (21%) | 207 (19%) | 433 (20%) |
| Jun-Aug 2022 | 241 (22%) | 307 (28%) | 548 (25%) |
| Sep-Nov 2022 | 359 (33%) | 334 (30%) | 693 (31%) |
| Dec 2022-Feb 2023 | 275 (25%) | 256 (23%) | 531 (24%) |
| <sup>1</sup>Includes only interviewees who consented and met all inclusion criteria. |  |  |  |

### Outcomes

We found no statistically significant impact of the intervention on the primary outcome of the proportion of ACTs sold to test-positive clients (Intervention: Control, Adj RR=0.85 [0.34 to 2.16]: 8.8% (49/558) in the control vs. 7.8% (49/626) in intervention arm). We also did not see any statistically significant impact on any of the other secondary outcomes (i.e., adherence to test result and appropriate case management); Intervention: Control, Adj RR=0.76 (0.36 to 1.62) and 0.67 (0.33 to 1.33), respectively (Table 2).

**Table 2:** Minimally adjusted and fully adjusted estimates of intervention effects for pre-specified study outcomes.

| Arm | Proportion | Risk Ratio (95% CI) |  | Risk Difference (95% CI) |  |
| --- | --- | --- | --- | --- | --- |
|  |  | Minimally | Fully | Minimally | Fully |
|  |  | Adjusted | Adjusted | Adjusted | Adjusted |
| Primary – ACTs sold to malaria test-positive clients |  |  |  |  |  |
| Control | 49/558 (8.8%) | — | — | — | — |
| Intervention | 49/626 (7.8%) | 0.85 | 0.79 | -0.01 | -0.02 |
|  |  | (0.34, 2.16) | (0.27, 2.33) | (-0.09, 0.07) | (-0.10, 0.07) |
| Secondary 1 -- Suspected malaria cases that receive a mRDT (testing uptake) |  |  |  |  |  |
| Control | 226/928 (24%) | — | — | — | — |
| Intervention | 217/958 (23%) | 0.92 | 0.95 | -0.01 | 0.01 |
|  |  | (0.47, 1.78) | (0.50, 1.81) | (-0.20, 0.18) | (-0.18, 0.20) |
| Secondary 2 – Malaria tested clients whose treatment adhered to test results |  |  |  |  |  |
| Control | 167/226 (74%) | — | — | — | — |
| Intervention | 118/217 (54%) | 0.76 | 0.77 | -0.17 | -0.17 |
|  |  | (0.36, 1.62) | (0.36, 1.64) | (-0.36, 0.01) | (-0.37, 0.03) |
| Secondary 3 – Suspected malaria cases that are managed appropriately |  |  |  |  |  |
| Control | 167/928 (18%) | — | — | — | — |
| Intervention | 118/958 (12%) | 0.67 | 0.66 | -0.05 | -0.06 |
|  |  | (0.33, 1.33) | (0.33, 1.31) | (-0.19, 0.10) | (-0.24, 0.11) |
| Secondary 4 – Untested clients taking ACT |  |  |  |  |  |
| Control | 462/836 (55%) | — | — | — | — |
| Intervention | 498/827 (60%) | 1.09 | 1.02 | 0.05 | 0.01 |
|  |  | (0.62, 1.90) | (0.60, 1.74) | (-0.03, 0.12) | (-0.08, 0.09) |

### Testing rates

Prior to the study, testing in the retail outlets was rarely offered. A survey performed prior to the start of the study found that only 12% (n=153) of outlets in the study area had mRDTs in stock during the time of the survey. Overall, during the study, 23% (443/1,886) of suspected cases (any client who was tested with an mRDT or was untested but purchased an antimalarial (AM) of any type) received an RDT at the outlet. We found no statistically significant impact of the intervention on testing rates (Intervention: Control, Adj RR=0.92 [0.47 to 1.78]; 23% (217/958) of suspected cases were tested in the intervention arm compared to 24% (226/928) in the control arm). Clients sometimes arrived with a test or prescription from a private laboratory or physician. In exit interviews, about 5% (94/1,886) of suspected cases came to the outlet with a diagnosis from another facility or laboratory. Few of these clients chose to retest (n=4). Of the clients that were tested in the outlet, 56% were female and 44% male. Of the clients that were not tested, 47% were female and 53% male. Most tests were done in the Sept-Nov period (37%), vs. 20-25% in the other three periods. No other discernable differences in wealth, education or age were observed when comparing those who were and were not tested (Table 3, Table S1).

**Table 3:**
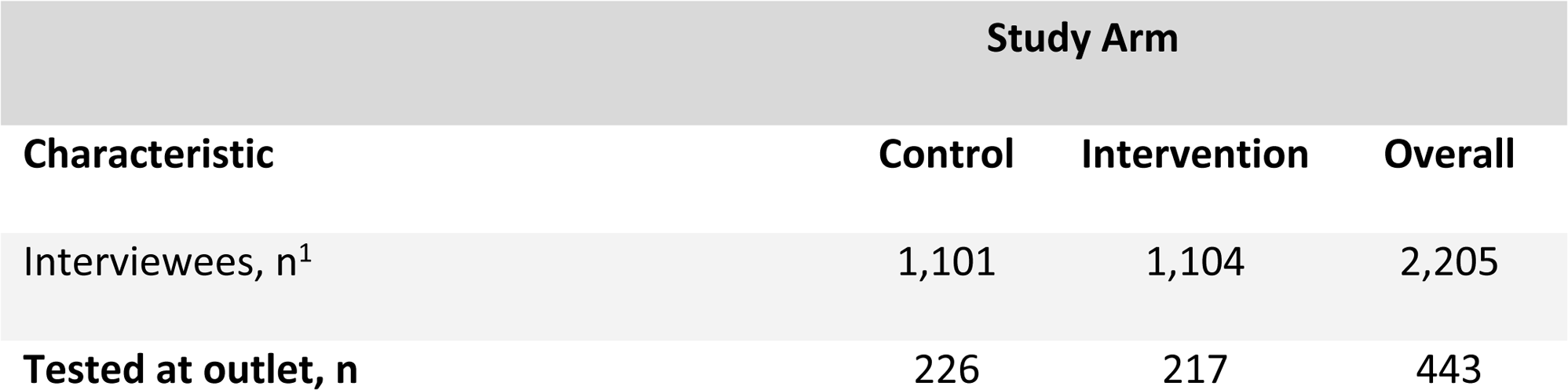
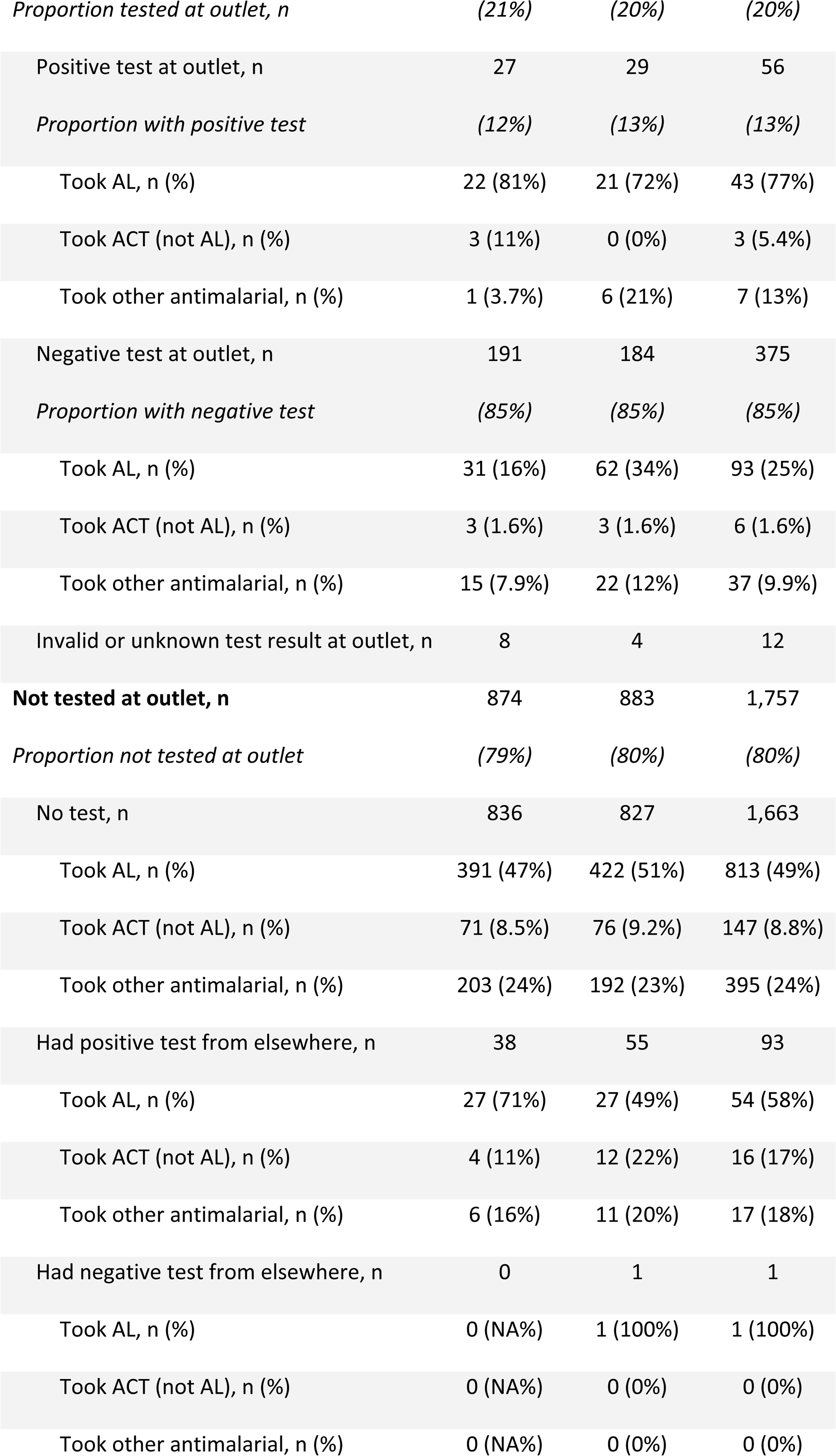
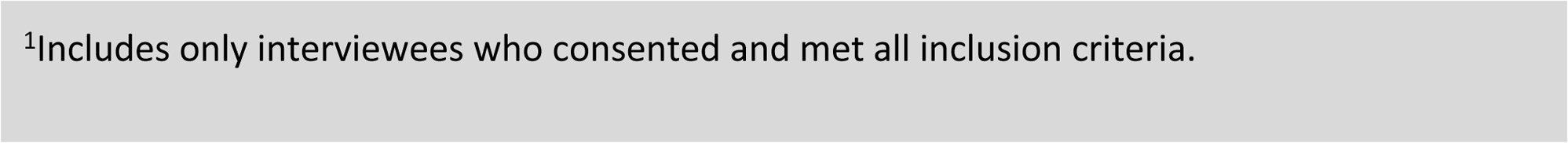
Testing and treatment decisions by arm.

| Characteristic | Study Arm |  |  |
| --- | --- | --- | --- |
|  | Control | Intervention | Overall |
| Interviewees, n<sup>1</sup> | 1,101 | 1,104 | 2,205 |
| Tested at outlet, n | 226 | 217 | 443 |
| *Proportion tested at outlet, n* | *(21%)* | *(20%)* | *(20%)* |
| Positive test at outlet, n | 27 | 29 | 56 |
| *Proportion with positive test* | *(12%)* | *(13%)* | *(13%)* |
| Took AL, n (%) | 22 (81%) | 21 (72%) | 43 (77%) |
| Took ACT (not AL), n (%) | 3 (11%) | 0 (0%) | 3 (5.4%) |
| Took other antimalarial, n (%) | 1 (3.7%) | 6 (21%) | 7 (13%) |
| Negative test at outlet, n | 191 | 184 | 375 |
| *Proportion with negative test* | *(85%)* | *(85%)* | *(85%)* |
| Took AL, n (%) | 31 (16%) | 62 (34%) | 93 (25%) |
| Took ACT (not AL), n (%) | 3 (1.6%) | 3 (1.6%) | 6 (1.6%) |
| Took other antimalarial, n (%) | 15 (7.9%) | 22 (12%) | 37 (9.9%) |
| Invalid or unknown test result at outlet, n | 8 | 4 | 12 |
| **Not tested at outlet, n** | **874** | **883** | **1,757** |
| *Proportion not tested at outlet* | *(79%)* | *(80%)* | *(80%)* |
| No test, n | 836 | 827 | 1,663 |
| Took AL, n (%) | 391 (47%) | 422 (51%) | 813 (49%) |
| Took ACT (not AL), n (%) | 71 (8.5%) | 76 (9.2%) | 147 (8.8%) |
| Took other antimalarial, n (%) | 203 (24%) | 192 (23%) | 395 (24%) |
| Had positive test from elsewhere, n | 38 | 55 | 93 |
| Took AL, n (%) | 27 (71%) | 27 (49%) | 54 (58%) |
| Took ACT (not AL), n (%) | 4 (11%) | 12 (22%) | 16 (17%) |
| Took other antimalarial, n (%) | 6 (16%) | 11 (20%) | 17 (18%) |
| Had negative test from elsewhere, n | 0 | 1 | 1 |
| Took AL, n (%) | 0 (NA%) | 1 (100%) | 1 (100%) |
| Took ACT (not AL), n (%) | 0 (NA%) | 0 (0%) | 0 (0%) |
| Took other antimalarial, n (%) | 0 (NA%) | 0 (0%) | 0 (0%) |

<sup>1</sup>Includes only interviewees who consented and met all inclusion criteria.

### ACT consumption and adherence

Across the study arms, we observed that testing seemed to have improved targeting of ACT. Of those testing positive, 82.1% (46/56) purchased an ACT, compared to 26.4% (99/375) of those testing negative. Amongst untested clients, 57.7% (960/1663) purchased an ACT. Overall, female clients had higher adherence to negative test results (i.e., tested negative with mRDT and not purchased any anti-malarial) than male clients (69% (146/213) vs. 57% (93/162)); male clients, in contrast, had higher adherence to positive results. (i.e., tested positive with mRDT and purchased ACT) 88% (23/26) vs. 77% (23/30). Of the seven women not purchasing an ACT (7/30), five purchased other antimalarials (Table 3, Table S2).

### Understanding the intervention

We found significant heterogeneity among participating outlets in RDT uptake. To understand the variability, we examined differences in testing and treatment at the cluster level. We found a handful of outlets were responsible for the vast majority of testing, with five outlets (out of 48) accounting for 41% of mRDTs administered. Testing rates ranged from as low as 0% of suspected cases to more than 85%. Although integration of testing into their business practices was highly heterogenous, the use of ACT or other antimalarial by untested clients was uniformly high with much less variation between clusters (Fig 2).

**Figure 2:**
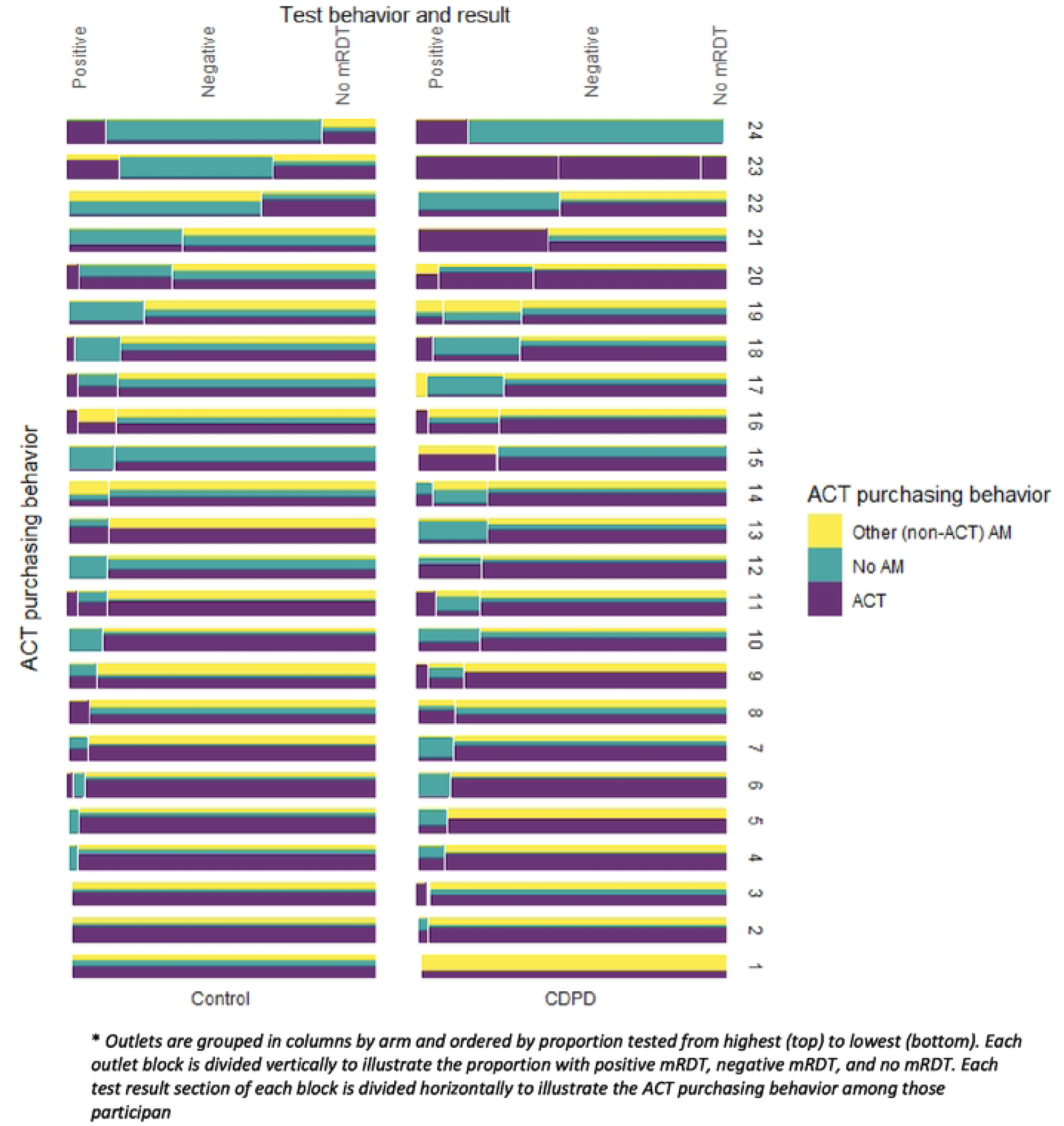
ACT and RDT uptake by outlet*.

The positivity rate of mRDTs conducted in the outlets was 13% as reported by customers exiting the outlets, with no difference between the intervention and control arm. The low testing rate, combined with a low positivity rate resulted in poor targeting of ACTs, with only 8.3% of all ACTs going to clients with confirmed malaria (Table 2, Table 3). Shops largely adhered to the recommended retail price for RDTs. Across arms, 85% (128/151) of clients confirmed they paid the RRP of 250 naira with only a small percentage paying more (5.3% or 8/151). Only a few clients that tested positive recalled what they paid for the ACT (28/56) and none of them reported receiving the ACT for free as intended by the intervention. Among those purchasing any ACTs, 50% reported paying >850 naira (263/527) with 29% of clients paying 650-850 naira (153/527). The mean price was 864 Naira with a median price of 700 Naira. No notable differences in ACT prices were observed across arms (Table 4).

**Table 4:** Summary of price variables among ACT purchasers.

| Characteristics | Control, N =<br>558 | Intervention, N =<br>626 | Overall, N =<br>1,184 |
| --- | --- | --- | --- |
| mRDT cost (Naira, categorized), n (%) |  |  |  |
| 0 | 0 (0%) | 5 (5.8%) | 5 (3.3%) |
| Don't know | 3 (4.6%) | 7 (8.1%) | 10 (6.6%) |
| >250 | 4 (6.2%) | 4 (4.7%) | 8 (5.3%) |
| 250 | 58 (89%) | 70 (81%) | 128 (85%) |
| Missing | 493 | 540 | 1,033 |
| ACT cost (Naira, categorized), n (%) |  |  |  |
| 0 | 0 (0%) | 0 (0%) | 0 (0%) |
| 1-649 | 33 (13%) | 32 (12%) | 65 (12%) |
| 650-850 | 73 (29%) | 80 (30%) | 153 (29%) |
| >850 | 131 (51%) | 132 (49%) | 263 (50%) |
| Don't know | 19 (7.4%) | 27 (10.0%) | 46 (8.7%) |
| Missing | 302 | 355 | 657 |
| ACT cost among test positive (Naira, categorized), n (%) |  |  |  |
| 0 | 0 (0%) | 0 (0%) | 0 (0%) |
| 1-649 | 7 (39%) | 1 (5.6%) | 8 (22%) |
| 650-850 | 2 (11%) | 1 (5.6%) | 3 (8.3%) |
| >850 | 7 (39%) | 10 (56%) | 17 (47%) |
| Don't know | 2 (11%) | 6 (33%) | 8 (22%) |
| Missing | 540 | 608 | 1,148 |
| ACT cost among test negative/untested (Naira, categorized), n (%) |  |  |  |
| 1-649 | 26 (11%) | 31 (12%) | 57 (12%) |
| 650-850 | 71 (30%) | 79 (31%) | 150 (31%) |
| >850 | 124 (52%) | 122 (48%) | 246 (50%) |
| Don't know | 17 (7.1%) | 21 (8.3%) | 38 (7.7%) |
| Missing | 320 | 373 | 693 |
| Why did you not pay anything for your ACT today?, n (%) |  |  |  |
| Charged to insurance | 0 (NA%) | 0 (NA%) | 0 (NA%) |
| Paid on credit | 0 (NA%) | 0 (NA%) | 0 (NA%) |
| The ACT was offered for free at the outlet | 0 (NA%) | 0 (NA%) | 0 (NA%) |
| Missing | 558 | 626 | 1,184 |
| Was there any discount for your ACT at the outlet today?, n (%) |  |  |  |
| Yes | 28 (11%) | 29 (10%) | 57 (11%) |
| No | 124 (48%) | 149 (54%) | 273 (51%) |
| Don't know | 106 (41%) | 99 (36%) | 205 (38%) |
| Missing | 300 | 349 | 649 |

Clients testing positive reported spending a higher total amount at the outlet-a median of 1,425 Naira (n=56) vs. 950 Naira for those testing negative (n= 375) and 850 Naira for those not testing (n=1757), respectively (Table 5).

**Table 5:** Total spent by test behavior and result.

| Characteristic | Positive, N =<br>56 | Negative, N<br>= 375 | No mRDT, N<br>= 1,757 | Overall |
| --- | --- | --- | --- | --- |
| How much did you spend at the pharmacy today? ('Don't know' as missing) |  |  |  |  |
| Mean (SD) | 1,534 (718) | 1,078 (728) | 1,026<br>(1,085) | 1,048<br>(1,034) |
| Median [IQR] | 1,425 [1,113,<br>2,000] | 950 [550,<br>1,350] | 850 [600,<br>1,200] | 900 [600,<br>1,300] |
| Missing | 2 | 77 | 107 | 186 |

Clients did not purchase injections in any significant proportion; 4.7% (6/127) of clients testing positive and 1.5% (25/1703) of clients untested. Just over a third (36%) of clients testing positive (20/56) purchased antibiotics; compared to 18% of those testing negative (66/375) and 29% (506/1,757) of untested clients (Table S3, Table S4).

## Discussion

There have been several studies that have shown the feasibility introducing mRDTs in PMRs, including in Nigeria, with free or low cost mRDTs offered to clients, generally resulting in higher testing uptake among clients compared to a control arm or baseline survey [20, 21,22]. In our study, we provided low cost RDTs to both the intervention and control arms, with an additional financial incentive for PPMVs in the intervention arm for each mRDT performed and reported through the study app, as well as a free ACT for clients receiving a positive mRDT result.

PMRs in both arms largely adhered to the recommended RDT price. We found approximately 23% of clients purchasing the mRDT. The additional incentive in the intervention arm, however, did not result in any additional impact on testing.

Similarly, findings from other studies [23, 24, 25], show that introducing mRDTs in PMRs improved ACT targeting with most clients procuring an ACT upon a positive result and a smaller proportion of clients procuring an ACT or another antimalarial with a negative result. However, instructing the PMRs in the intervention arm, to give the ACT for free to clients conditional on a positive result, like we did in our study, did not have any additional impact on testing or targeting.

Study results in western Kenya, in which we implemented a similar trial, also found little impact of the intervention on testing uptake or targeting of ACTs. Across both sites, we found testing uptake highly heterogeneous among PMRs with many clients testing negative purchasing antimalarials and most clients foregoing testing altogether while purchasing ACTs or other antimalarials. Clients testing positive in both countries also spent more (and purchased more drugs) compared to untested patients or patients testing negative. These results were found in both arms, with clients testing positive in the intervention arm indicating they did not receive a discount or receive the ACT for free, as the intervention intended.

There were, however, notable differences between the two study sites. In Kenya, we found a much higher proportion of clients visiting the participating PMRs with malaria like symptoms compared to Nigeria (i.e., eligible clients made up 51% of clients in Kenya, vs. 19% in Lagos and 82% of ineligible clients in Lagos responded not experiencing malaria symptoms vs. 11% in Kenya). This higher proportion was likely due to higher malaria prevalence in the Kenyan study site and may have contributed to the much higher testing and positivity rate found in Kenya compared to Lagos (i.e., 49% and 35% compared to 23% and 13% in Kenya and Nigeria, respectively). As a result, a much larger proportion of ACTs were dispensed to clients testing positive in Kenya compared to Nigeria (27% vs. 8%, respectively). Clients also purchased different types of drugs. In Kenya, a sizable proportion of clients testing positive purchased injections (22%) instead of or in addition to an ACT compared to Nigeria (4.7%). Clients in Kenya also were more likely to purchase ACT types other than AL compared to Nigeria.

The varying results between the two sites in testing uptake and treatment choice may reflect differences in malaria prevalence, client preferences and their socio-economic status, or market conditions, among other factors. Yet, the PMR-oriented financial incentive approach we took to increase RDT uptake and lower the price of the ACTs to the client, conditional on the result of the mRDT, did not have an impact on client or provider behavior in either setting. The shop attendants may not have understood or been aware of the intervention, and profit incentives may have constrained behavior change, with shop attendants charging for ACTs for RDT-positive clients in the intervention arm [26]. Suggestions from participating PMRs following the study, solicited through group discussions, to increase test uptake and adherence included further lowering the cost of the mRDT and ACT, building trust with clients around the (negative) RDT result (i.e., clients often come in believing they have malaria) and alleviating concerns that clients may see getting the test as an inconvenience. Results from a recent three arm randomized control trial by Omale *et al* (2021) in Nigeria found that sensitizing caregivers through social group meetings around the need for testing had a significant impact on testing uptake for under 5s in PPMVs, with no significant difference between the social group arm and the arm that combined the social group meetings with provider trainings [27], suggesting that educating clients and caregivers, promoting RDT use, and advocating for adherence could further improve outcomes.

Efforts to increase use of RDTs in PMRs and adherence to test results should not only align with business practices to make RDTs and ACTs available and affordable but include demand-side community mobilization interventions, that could sensitize and empower patients to be sound decision makers around malaria testing and drug choice.

## Conclusions

Introduction of low cost RDTs to PMRs appears to improve targeting of ACTs among tested clients. However, providing financial incentives to PMRs to increase testing uptake and adherence may not result in additional improvements if the incentives are not well-understood or well-aligned with existing business practices. In addition to interventions that can lower the out-of-pocket cost of RDTs and ACTs for clients in PMRs, communities at risk should be sensitized to the benefits of testing in PMRs.

## Data Availability

All relevant data is already provided as part of the submitted article.

